# Cognitive function among former professional male soccer players – the HEADING study

**DOI:** 10.1101/2024.03.26.24304885

**Authors:** Valentina Gallo, Giulia Seghezzo, Ioannis Basinas, Elizabeth Williamson, Yvonne van Hoecke, Donna Davoren, Simon Kemp, Saba Mian, Sinead Langan, Henrik Zetterberg, Danielle Pearce, John W. Cherrie, Damien M McElvenny, Neil Pearce

## Abstract

**Background:** The HEalth and Ageing Data IN the Game of football (HEADING) study assessed the associations between exposure to heading a football, other impacts to the head, and concussions, with cognitive function, among former professional soccer players in England.

**Methods:** Recruitment of former male professional soccer players aged 50+ years was conducted through the English Professional Footballers’ Association. Cumulative exposure to heading and other impacts to the head was estimated from the playing history questionnaire. Concussion was self-reported and assessed with the BRAIN-Q tool. The primary outcome was cognitive function measured with the Preclinical Alzheimer Cognitive Composite (PACC).

**Findings:** Data for a total of 199 males were available for analysis. No overall association was found between heading and/or other impacts to the head and cognitive function. Amongst forwards only, those in the fourth (highest) quartile of exposure to other impacts to the head had a significantly lower PACC score compared to those in the first quartile (• = -0.65, 95% C.I. -1.23, -0.07). For all players, self-reported concussions were associated with slightly lower cognitive function (• = -0.01; 95% C.I. -0.01, -0.001).

**Interpretation:** This study generally does not support an association between exposure to heading a football and poorer cognitive function among former male professional players. However, an association between the number of concussions sustained and poorer cognitive function was present, although the effect size is relatively small.

**Funding:** This study was funded with a grant of the Drake Foundation to the London School of Hygiene and Tropical Medicine.

**Research in context:** *Evidence before this study:* There is increasing evidence for an association between sport-related concussion and poorer cognitive function later in life, in former athletes. In soccer, heading the ball is a common event, particularly for outfield players, and involves repetitive sub-concussive impacts. The association between exposure to heading and cognitive function later in life is not consistent in the literature.

*Added value of this study:* This study is the first to provide appropriately modelled exposure estimates of both heading the ball and other impacts to the head among soccer players. Our findings do not support an association between heading and poorer cognitive function later in life. However they do suggest an association between concussion and poorer cognitive function among soccer players, although the size of the effect is relatively small. Other impacts to the head, such as head-to-head collisions, that did not result in the clinical features of concussions were also shown to be associated with poorer cognitive function, but only among forwards.

*Implication of the available evidence:* These findings support a continuous focus on the prevention of concussion in all sports, including soccer, whilst the consequences of repeated exposure to head impacts need to be further explored. Methodologically, it would be advisable to harmonise the tools developed to estimate cumulative exposures to heading, to other head impacts, and to sport-related concussion, to increase comparability of results across studies, thus enabling a sound synthesis of the evidence.

## Introduction

Evidence for an association between concussion and poorer cognitive function among participants of contact sports, i.e. where players deliberately or accidentally hit or collide with each other or with inanimate objects, is mounting^1^. Despite not being regarded as a contact sport, in association football (soccer) player-to-player contact is unavoidable, including the potential for acute head injury/concussion^2^. Moreover, outfield players not only play with their feet, but they also head the ball, which can reach speeds of 35m/s^3^, leading to exposure to repetitive sub-concussive head impacts^4^. Therefore, concern over the long-term neurological health and cognitive function of soccer players is increasing.

A few studies have recently addressed this concern, using external (soccer players vs. the general population) or internal (high vs. low exposed to head impacts among soccer players) comparisons^1^. In general, soccer players seem to be at increased risk of cognitive decline compared to the general population. A record-linkage analysis of Scottish professional soccer players found a five-fold increase in mortality for Alzheimer’s disease among outfield soccer players (but a weaker association in goalkeepers) compared to the general population (hazard ratio (HR) 5.07; 95% CI, 2.92 to 8.82)^5^. This relative risk was reduced, but remained statistically significant, after also including incident cases identified through clinical records (HR 3.59; 95% CI 1.81 to 4.39)^6^. Another analysis of a sample from the same population found an increased prevalence of neurodegenerative diseases among former soccer players compared to controls^7^. Similarly, an increased risk of Alzheimer disease was reported in a large, nationwide cohort study of Swedish soccer players compared to the general population (HR 1.62; 95% C.I. 1.47 to 1.78)^8^. Mortality from dementia was found to be higher among French soccer players compared to the general population (SMR: 3.38; 95% CI: 2.49 to 4.46) in another large, nationwide study^9^.

The results are less consistent when internal comparisons are conducted. Among Scottish players the risk of dying from dementia increased with increasing length of career^6^, and varied greatly by position played, supporting a possible role of cumulative heading. Conversely, another smaller study conducted in England and Scotland did not find an association between position or length of career and mild cognitive impairment among former soccer players^10^. Two other studies recruiting former soccer players from the United Kingdom, with unclear population overlap, found associations between frequency of heading and cognitive impairment^11,12^. Conversely, no difference in cognitive performance was found comparing current players and population controls, either among male Brazilian professional soccer players^13^, or among female players, in Europe^14^. A systematic review of the short-term effects of heading the ball among soccer players on neurocognitive performance found no statistically significant effects^15^. Finally, a human experimental study of controlled heading from goalkeeper kickouts found no cerebrospinal fluid biomarker evidence of neuronal or astroglial injury from the head impacts^16^.

The aim of the HEalth and Ageing Data IN the Game of football (HEADING) study was to assess the associations between heading, other impacts to the head, and concussion - separately and in combination - with cognitive function later in life, among former professional soccer players. To do so, a method for assessing exposure to heading and other impacts to the head, based on validated questionnaires and statistical modelling, was developed.

## Methods

The HEADING study is a cross-sectional study with a design similar to the previously published BRAIN study which assessed the association between self-reported concussion and cognitive function among former elite rugby players^17,18^. In the HEADING study, an additional component assessing the exposure to heading and other impacts to the head, was included^19^. The study recruited former professional male soccer players aged 50+ between July 2019 and December 2021. Current and former members of the English Professional Footballers’ Association (PFA)^20^ were invited to take part into the study by the PFA by post and email (N=1,569), and were sent a remainder by email only (N=192). It was estimated that by recruiting 150 former players, considering that the standard deviations (SDs) of the psychometric tests are in the range of 8%–15% of the absolute value, the study would have more than 95% power to detect a 10% difference, and 80% power to detect a 7% difference in psychometric test scores between exposed and non-exposed participants^17^.

Initially, those who agreed to participate were assessed in a clinic in London (N=9), in Manchester (N=14), or at home (N=20). After the COVID-19 restrictions were imposed, assessment was moved online (N=99), after we had conducted a study to confirm the validity of this approach for assessing the Preclinical Alzheimer Cognitive Composite (PACC)^21^. The assessment included questions on soccer playing career, occupational history, lifestyle factors, cognitive ability and self-reported history of concussion^17^. At the end of the interview, participants were asked to provide detailed information about their history of heading, other impacts to the head, and concussion while playing and training before, during and after their professional career. Participants who were cognitively impaired were assessed with the assistance of a close family member. All participants signed an informed consent. The study was approved by the Ethical Committee of the London School of Hygiene and Tropical Medicine (EC/16282). The study protocol can be accessed at https://www.lshtm.ac.uk/research/centres-projects-groups/heading-study.

### Exposure definition and assessment

Concussion was defined according to the Berlin consensus as an alteration in brain function, caused by an external force, with or without loss of consciousness^17,22^. This definition was recently revised, without substantial change for the scope of the study^23^. Headings were defined as self-reported intentional impacts between the head and the ball^19^. Non-heading impacts were defined as head-to-head collisions and direct blows to the head which did not result in clinical symptoms of a concussion^19,24,25^.

Cumulative exposure to heading and other impacts to the head was estimated from the participants’ playing history obtained from dedicated questionnaire developed with extensive piloting, validation and input from the PFA, the Football Association (FA), and the coaching staff of Sheffield United Football Club^19^. Cumulative exposure was calculated as the number of episodes (exposure to heading impacts, and other impacts to the head) over a playing career, including juvenile, and post retirement^26^. Modelled exposure estimates were used to minimise the presence of potential measurement errors due to recall^27^ and to allow the assignment of exposure estimates to participants with missing information (n=12). The average frequency of heading and other impacts to the head per match, or training event by position, playing level, and decade of play were estimated from a linear mixed effect regression model with player as the random effect^19^. The model performance and the validity of the estimates were evaluated internally against the results from a subset of records not used in the development of the model and externally by comparisons with the available literature.

Information on concussions was collected using the BRAIN-Q tool^28^, designed specifically for sport-related concussions^18^, and validated in its telephone version BRAIN-Qt^28^. The total number of soccer-related concussions, was categorised into “no concussion”, “low concussion (1 or 2)”, and “high concussion (3+)”. Length of professional playing career was also used as a proxy for cumulative exposure to all impacts.

### Assessment of cognitive function

The primary outcome measure was an adapted version of the Alzheimer Disease Cooperative Study – Preclinical Alzheimer Cognitive Composite (ADCS-PACC)^29^. The PACC includes: 1) The Total Recall score from the Free and Cued Selective Reminding Test (FCSRT) (0-48 words)^30^; 2) The Delayed Recall score on the Logical Memory-II, subtest from the Wechsler Memory Scale (0-25 story units)^29^; 3) The Digit Symbol Substitution Test score from the Wechsler Adult Intelligence Scale–Revised (0-93 symbols)^29^; 4) The Mini-Mental State Examination (MMSE) (0-30 points)^29^. As for the BRAIN study, the FCSRT was replaced with the total score of the 12-item face-name test (FNAME-12), 0-96 points^31^, which is similar in terms of testing immediate and delayed recall^32^, as it showed evidence of convergent validity with the established paired associative memory task, FCSRT (Pearson *r*=0.32, p<0.05)^31^. The test scores of the four components of the PACC were standardized to create z-scores based on the overall HEADING Study sample and averaged. At least three tests were required to create a PACC score, one of them being the MMSE^29^.

The COVID-19 restrictions came into place when 43 participants had already been assessed in person; 28 (65%) of these were recalled and reassessed online in order to estimate the validity of remote assessment using the PACC. This validation^21^ indicated that there was a good correlation between the two methods (Pearson correlation coefficient between the live and telephonic assessment 0.82 (95% CI 0.66, 0.98)). Thus, the rest of the sample was assessed online.

### Covariates and potential confounders

Potential confounding variables and their categorisation were pre-specified in the analysis plan and – where possible –were aligned with those of the BRAIN study^17^. Age in years was recorded at the time of the assessment. Highest educational qualification was classified into: primary school, GCSE or equivalent, A-level or above. Tobacco smoking and alcohol drinking were classified as current/former/never. Diagnoses of hypertension and diabetes were recorded. Given the online nature of the assessment for part of the sample, it was not possible to record height and weight. Ethnicity was self-reported and classified as white, Black, or mixed. Soccer positions were classified as defender, forward, goalkeeper, and midfielder or utility player.

### Statistical analysis

Missing values for cognitive tests, exposure variables, and covariates were imputed using multiple imputation by chained equations (MICE), i.e., using a fully conditional specification^33^, creating ten imputed datasets^34^. Individual imputation models used logistic regression for binary variables, ordinal logistic regression for ordered categorical variables, and multinomial regression for unordered categorical variables. For continuous variables with missing values, predictive mean matching was used, with 5 nearest neighbours, to address potential issues with non-normality^34^.

Linear regression models were conducted for the PACC score, with quartiles of distribution of cumulative exposure to heading, other impacts to the head, the total number of concussions and its grouping in categories, and length of playing career in years. The analyses were adjusted for pre-specified potential confounders: age (continuous), education, smoking, alcohol, medical history, soccer playing position (only for concussion), and study assessor. A possible interaction between concussion history and age in 10-years age bands was assessed. Sensitivity analyses were conducted to test the main model after removing the imputed values, after removing study assessor, and replacing modelled exposure to heading with self-reported exposures.

## Results

Of the 1,761 invited, 212 (12%) agreed to participate, and 199 (94%) of these were interviewed. Of the 199 interviewed, complete data were available for 142 (71%) participants. During the initial COVID-19 restrictions, some participants were just assessed for the basic demographic information and playing history and were later recalled (once we had conducted the validation study) for the cognitive function testing, but we were not able to successfully recall all of them; similarly some other participants did not provide their playing history but completed all other components. Thus, the final sample for analysis was 199 after MICE imputations were performed. The participants’ demographic and playing characteristics by quartiles of cumulative heading and by categories of concussions are shown in Table 1 and Supplementary Table 1, respectively. The participants had a median age (p25-p75) of 63 years (58-71), and only 41 (29%) of them achieved an A-level or above. The mean (SD) length of their playing career was 17.5 (5.9) years; with 35% being defenders, 18% forwards, 3% goalkeepers, 21% midfielders, and 23% utility players. The findings for the individual tests of the PACC are reported in Supplementary Table 2; PACC scores by quartiles of distribution of heading and concussion categories are shown in Figure 1.

**Table 1:**
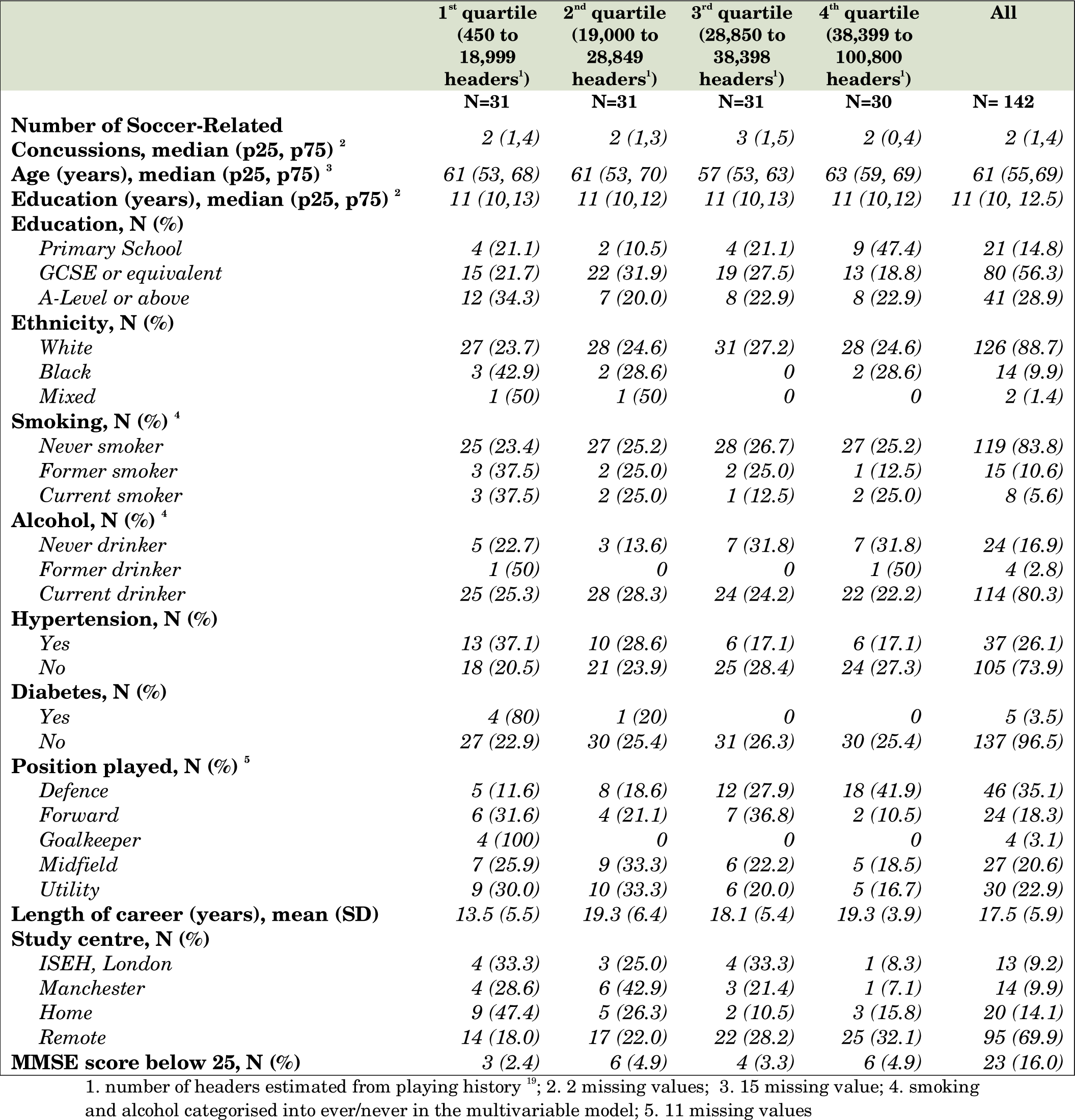
Characteristics of study participants by quartiles of distribution of cumulative numbers of modelled heading

|  | **1<sup>st</sup> quartile**<br>**(450 to**<br>**18,999**<br>**headers<sup>1</sup>)**<br>**N=31** | **2<sup>nd</sup> quartile**<br>**(19,000 to**<br>**28,849**<br>**headers<sup>1</sup>)**<br>**N=31** | **3<sup>rd</sup> quartile**<br>**(28,850 to**<br>**38,398**<br>**headers<sup>1</sup>)**<br>**N=31** | **4<sup>th</sup> quartile**<br>**(38,399 to**<br>**100,800**<br>**headers<sup>1</sup>)**<br>**N=30** | **All**<br>**N= 142** |
| --- | --- | --- | --- | --- | --- |
| **Number of Soccer-Related Concussions, median (p25, p75) <sup>2</sup>** | 2 (1,4) | 2 (1,3) | 3 (1,5) | 2 (0,4) | 2 (1,4) |
| **Age (years), median (p25, p75) <sup>3</sup>** | 61 (53, 68) | 61 (53, 70) | 57 (53, 63) | 63 (59, 69) | 61 (55,69) |
| **Education (years), median (p25, p75) <sup>2</sup>** | 11 (10,13) | 11 (10,12) | 11 (10,13) | 11 (10,12) | 11 (10, 12.5) |
| **Education, N (%)** |  |  |  |  |  |
| *Primary School* | 4 (21.1) | 2 (10.5) | 4 (21.1) | 9 (47.4) | 21 (14.8) |
| *GCSE or equivalent* | 15 (21.7) | 22 (31.9) | 19 (27.5) | 13 (18.8) | 80 (56.3) |
| *A-Level or above* | 12 (34.3) | 7 (20.0) | 8 (22.9) | 8 (22.9) | 41 (28.9) |
| **Ethnicity, N (%)** |  |  |  |  |  |
| *White* | 27 (23.7) | 28 (24.6) | 31 (27.2) | 28 (24.6) | 126 (88.7) |
| *Black* | 3 (42.9) | 2 (28.6) | 0 | 2 (28.6) | 14 (9.9) |
| *Mixed* | 1 (50) | 1 (50) | 0 | 0 | 2 (1.4) |
| **Smoking, N (%) <sup>4</sup>** |  |  |  |  |  |
| *Never smoker* | 25 (23.4) | 27 (25.2) | 28 (26.7) | 27 (25.2) | 119 (83.8) |
| *Former smoker* | 3 (37.5) | 2 (25.0) | 2 (25.0) | 1 (12.5) | 15 (10.6) |
| *Current smoker* | 3 (37.5) | 2 (25.0) | 1 (12.5) | 2 (25.0) | 8 (5.6) |
| **Alcohol, N (%) <sup>4</sup>** |  |  |  |  |  |
| *Never drinker* | 5 (22.7) | 3 (13.6) | 7 (31.8) | 7 (31.8) | 24 (16.9) |
| *Former drinker* | 1 (50) | 0 | 0 | 1 (50) | 4 (2.8) |
| *Current drinker* | 25 (25.3) | 28 (28.3) | 24 (24.2) | 22 (22.2) | 114 (80.3) |
| **Hypertension, N (%)** |  |  |  |  |  |
| *Yes* | 13 (37.1) | 10 (28.6) | 6 (17.1) | 6 (17.1) | 37 (26.1) |
| *No* | 18 (20.5) | 21 (23.9) | 25 (28.4) | 24 (27.3) | 105 (73.9) |
| **Diabetes, N (%)** |  |  |  |  |  |
| *Yes* | 4 (80) | 1 (20) | 0 | 0 | 5 (3.5) |
| *No* | 27 (22.9) | 30 (25.4) | 31 (26.3) | 30 (25.4) | 137 (96.5) |
| **Position played, N (%) <sup>5</sup>** |  |  |  |  |  |
| *Defence* | 5 (11.6) | 8 (18.6) | 12 (27.9) | 18 (41.9) | 46 (35.1) |
| *Forward* | 6 (31.6) | 4 (21.1) | 7 (36.8) | 2 (10.5) | 24 (18.3) |
| *Goalkeeper* | 4 (100) | 0 | 0 | 0 | 4 (3.1) |
| *Midfield* | 7 (25.9) | 9 (33.3) | 6 (22.2) | 5 (18.5) | 27 (20.6) |
| *Utility* | 9 (30.0) | 10 (33.3) | 6 (20.0) | 5 (16.7) | 30 (22.9) |
| **Length of career (years), mean (SD)** | 13.5 (5.5) | 19.3 (6.4) | 18.1 (5.4) | 19.3 (3.9) | 17.5 (5.9) |
| **Study centre, N (%)** |  |  |  |  |  |
| *ISEH, London* | 4 (33.3) | 3 (25.0) | 4 (33.3) | 1 (8.3) | 13 (9.2) |
| *Manchester* | 4 (28.6) | 6 (42.9) | 3 (21.4) | 1 (7.1) | 14 (9.9) |
| *Home* | 9 (47.4) | 5 (26.3) | 2 (10.5) | 3 (15.8) | 20 (14.1) |
| *Remote* | 14 (18.0) | 17 (22.0) | 22 (28.2) | 25 (32.1) | 95 (69.9) |
| **MMSE score below 25, N (%)** | 3 (2.4) | 6 (4.9) | 4 (3.3) | 6 (4.9) | 23 (16.0) |

1. number of headers estimated from playing history <sup>19</sup>; 2. 2 missing values; 3. 15 missing value; 4. smoking and alcohol categorised into ever/never in the multivariable model; 5. 11 missing values

**FIGURE 1:**
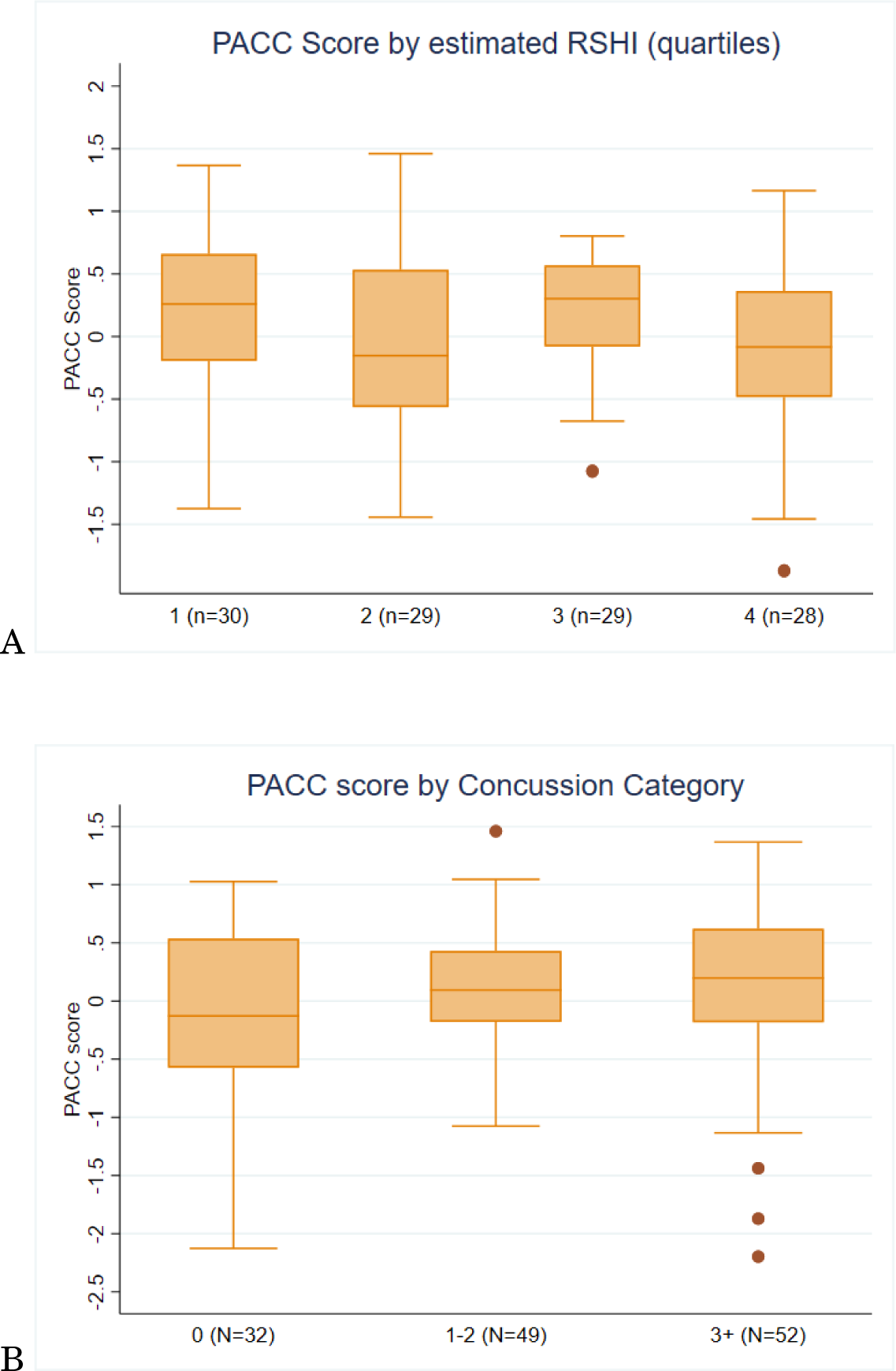
DISTRIBUTION OF PACC SCORES BY QUARTILES OF DISTRIBUTION OF ESTIMATED CUMULATIVE HEADING (A) AND BY CONCUSSION CATEGORIES (B)

The cumulative lifetime number of headers ranged between 415 and 100,785, with a median (p25-p75) of 28,842 (18,876, 38,397). Heading was associated with length of career (p<0.001) as well as position played (p=0.002) with 42% of the defenders in the highest quartile of heading, and only 19% of the midfielders, 11% of the forwards and none of the goalkeepers in the same category. Overall, little or no association was found between cumulative heading exposure and cognitive function as measured by the PACC, despite an apparent dose-response relationship of decreasing cognitive function with increasing quartiles of heading exposure in unadjusted models (Table 2). No significant differences were found in the associations between heading variables and PACC across playing positions (Table 3).

**Table 2:** age-adjusted and fully adjusted beta coefficinets coming from linear regression models investigating the association between estimated exposure to heading, other impacts to the head, and soccer-related concussions with the PACC score among former professional soccer players

|  | N | Age-adjusted Coefficient<br>(95% CI) | Fully adjusted Coefficient<br>(95% CI) | Mutually adjusted<br>Coefficients<sup>2</sup> (95% C.I.) |
| --- | --- | --- | --- | --- |
| **Heading <sup>1</sup>** |  |  |  |  |
| 1<sup>st</sup> quartile | 48 | Reference | Reference | Reference |
| 2<sup>nd</sup> quartile | 52 | -0.10 (-0.40, 0.19) | -0.13 (-0.46, 0.18) | -0.13 (-0.44, 0.18) |
| 3<sup>rd</sup> quartile | 50 | -0.17 (-0.45, 0.11) | -0.12 (-0.44, 0.20) | -0.14 (-0.42, 0.15) |
| 4<sup>th</sup> quartile | 50 | -0.21 (-0.53, 0.10) | -0.07 (-0.39, 0.25) | -0.09 (-0.39, 0.22) |
| p-trend |  | 0.508 | 0.811 | 0.777 |
| Number of headers (continuous, per 1,000 episodes) | 199 | -0.005 (-0.012, 0.001) | -0.002 (-0.008, 0.004) | -0.001 (-0.008, 0.005) |
| **Other impacts to the head <sup>1</sup>** |  |  |  |  |
| 1<sup>st</sup> quartile | 39 | Reference | Reference | Reference |
| 2<sup>nd</sup> quartile | 51 | 0.05 (-0.37, 0.47) | 0.01 (-0.37, 0.38) | 0.02 (-0.35, 0.40) |
| 3<sup>rd</sup> quartile | 55 | 0.11 (-0.27, 0.48) | 0.07 (-0.25, 0.40) | 0.08 (-0.24, 0.41) |
| 4<sup>th</sup> quartile | 54 | 0.04 (-0.30, 0.38) | -0.02 (-0.35, 0.32) | 0.03 (-0.31, 0.36) |
| p-trend |  | 0.931 | 0.922 | 0.951 |
| Number of other impacts the head (continuous) | 199 | -0.0002 (-0.001, 0.001) | -0.0001 (-0.001, 0.001) | -0.00002 (-0.001, 0.001) |
| **Soccer-related Concussions** |  |  |  |  |
| No Concussions | 28 | Reference | Reference | Reference |
| Low Concussions (1-2) | 41 | 0.07 (-0.18, 0.32) | 0.09 (-0.16, 0.34) | 0.09 (-0.17, 0.35) |
| High Concussions (3+) | 47 | -0.08 (-0.33, 0.17) | -0.09 (-0.32, 0.01) | -0.08 (-0.32, 0.16) |
| p-trend |  | 0.410 | 0.274 | 0.322 |
| Number of concussions (continuous) | 199 | **-0.01 (-0.03, -0.001)** | **-0.01 (-0.02, -0.002)** | **-0.01 (-0.02, -0.001)** |
| **Length of career, years** | 199 | -0.004 (-0.02, 0.01) | -0.003 (-0.02, 0.01) | -0.003 (-0.02, 0.01) |

1. number of headers and other impacts to the head estimated from playing history <sup>19</sup>, the number of players in each quartiles are average across imputations; 2. Mutually adjusted models include the three categorical exposure metrics (quartiles of number of headers and of other impacts to the head) and categories of concussion or the three continuous exposure metrics (number of headers, number of other impacts to the head, and number of concussions) in the same model. Length of career is adjusted for the three continuous metrics.

**Table 3:** Regression coefficients and relative 95% C.I. from fully adjusted models (not imputed) of the association between exposure to heading, other impacts to the head, and soccer-related concussions (in categories and as continuous variables) and length of career with PACC scores, by position played

|  | N=50 | Defenders & goalkeepers<br>Fully adjusted<br>Coefficient (95% CI) | N= 24 | Forward<br>Fully adjusted<br>Coefficients (95% C.I.) | N=27 | Middle fielders<br>Fully adjusted<br>coefficients (95% C.I.) | N=30 | Utility players<br>Fully adjusted<br>coefficients (95% C.I.) |
| --- | --- | --- | --- | --- | --- | --- | --- | --- |
| **Heading<sup>1</sup>** |  |  |  |  |  |  |  |  |
| 1<sup>st</sup> quartile | 8 | Refence | 2 | Reference | 5 | Reference | 9 | Reference |
| 2<sup>nd</sup> quartile | 8 | 0.15 (-0.36, 0.66) | 4 | -0.88 (-2.65, 0.88) | 10 | -0.09 (-0.97, 0.79) | 7 | 0.13 (-0.62, 0.89) |
| 3<sup>rd</sup> quartile | 13 | 0.20 (-0.24, 0.63) | 6 | -0.76 (-1.99, 0.48) | 3 | -0.15 (-1.38, 1.68) | 7 | -0.17 (-0.97, 0.53) |
| 4<sup>th</sup> quartile | 8 | -0.27 (-0.82, 0.29) | 5 | -0.65 (-2.14, 0.83) | 1 | 0.66 (-1.32, 2.64) | 4 | 0.06 (-0.90, 1.02) |
|  | p-trend | 0.244 | p-trend | 0.471 |  | 0.800 |  | 0.877 |
| Number of headers<br>(continuous, per 1,000<br>episodes) | 38 | -0.009 (-0.02, 0.001) | 18 | -0.01 (-0.05, 0.03) | 19 | 0.001 (-0.03, 0.03) | 27 | -0.01 (-0.03, 0.01) |
| **Other impacts to the head<sup>1</sup>** |  |  |  |  |  |  |  |  |
| 1<sup>st</sup> quartile | 12 | Reference | 7 | Reference | 0 | Reference | 3 | Reference |
| 2<sup>nd</sup> quartile | 11 | 0.40 (-0.05, 0.85) | 6 | -0.10 (-1.14, 0.94) | 5 | - | 6 | -1.16 (2.40, 0.07) |
| 3<sup>rd</sup> quartile | 11 | 0.30 (-0.15, 0.75) | 3 | -0.03 (-1.17, 1.11) | 8 | -0.23 (-1.34, 0.87) | 8 | -0.72 (-1.83, 0.38) |
| 4<sup>th</sup> quartile | 7 | -0.18 (-0.55, 0.51) | 7 | -0.65 (-1.23, -0.07) | 6 | -0.60 (-1.61, 0.48) | 10 | -0.82 (-1.97, 0.32) |
|  | p-trend | 0.203 |  | 0.138 |  | 0.448 |  | 0.281 |
| Number of other impacts to<br>the head (continuous) | 41 | -0.001 (-0.004, 0.002) | 23 | -0.003 (-0.007, -0.0002) | 19 | -0.001 (-0.007, 0.005) | 27 | 0.0003 (-0.002, 0.003) |
| **Soccer-related Concussions** |  |  |  |  |  |  |  |  |
| No Concussions | 7 | Reference | 7 | Reference | 5 | Reference | 7 | Reference |
| Low Concussions (1-2) | 15 | -0.08 (-0.65, 0.49) | 8 | 0.54 (-0.17, 1.24) | 7 | 0.10 (-1.09, 1.28) | 10 | -0.14 (0.72, 0.44) |
| High Concussions (3+) | 19 | -0.14 (0.66, 0.37) | 8 | -0.18 (-0.86, 0.50) | 7 | -0.18 (-1.30, 0.94) | 9 | -0.31 (-0.98, 0.35) |
|  | p-trend | 0.846 |  | 0.06 |  | 0.850 |  | 0.597 |
| Number of concussions<br>(continuous) | 41 | -0.02 (-0.04, -0.001) | 23 | -0.01 (-0.04, 0.01) | 19 | -0.02 (-0.2, 0.16) | 26 | -0.05 (-0.14, 0.03) |
| **Length of career, years** | 41 | -0.03 (-0.07, -0.001) | 23 | 0.02 (-0.03, 0.06) | 19 | -0.01 (-0.11, 0.08) | 27 | 0.002 (-0.04, 0.04) |

1. number of headers and other impacts to the head estimated from playing history<sup>19</sup>

The cumulative number of impacts to the head (including both head-to-head collisions and blows to head) ranged from 0 to 503 in the sample, with a median (p25-p75) of 64 (26-120). These estimated head impacts were marginally associated with position played (p=0.06) but not with length of career (p=0.29). Overall, no association was found between estimated other head impacts and cognitive function as measured by the PACC (Table 2). However, players in the fourth quartiles of exposure to other head impacts had a significantly lower PACC score compared to those in the first quartile (• = -0.65, 95% C.I. -1.23, -0.07) among forwards only. In the same playing position group, the number of head impacts was negatively associated with PACC scores (• = -0.003, 95% C.I. -0.007, - 0.0002) (Table 3).

A total of 107 (76%) respondents reported at least one soccer-related concussion. Among the concussed, the number of lifetime concussions varied between one and 60, with a median (p25-p75) of 2 (1-4). The number of soccer-related concussions was marginally associated with position played (p=0.09), but not with length of career (p=0.54). The cumulative number of self-reported soccer-related concussions was associated with lower cognitive function as measured by the PACC both in fully adjusted models (• = -0.01; 95% C.I. -0.01, -0.001) and in models additionally adjusting for heading exposure (• = - 0.03, 95% CI (−0.03, -0.001)). When analysed by playing position, the observed number of concussions (continuous) was associated with lower PACC scores among defenders and goalkeepers only (• = -0.02, 95% C.I. -0.04, -0.001) (Table 3). Career length was not associated with measures of cognitive function, except among defenders and goalkeepers (• = -0.03, 95% C.I. -0.07, -0.001) (Table 3).

The sensitivity analysis conducted after excluding imputed values (Supplementary Table 5) and after removing study assessor from the model (results not shown) yielded virtually unchanged results.

## Discussion

This is the first study reporting the association of cognitive function with heading a football - based on appropriate modelled exposures estimates - together with other impacts to the head among former male professional soccer players. We do not find a clear association between exposure to heading and cognitive function later in life. However, stronger forms of impacts to the head – those associated with higher levels of acceleration compared to the heading^26^ – were found to be associated with poorer cognitive function, albeit among forwards only. Soccer-related concussions were found to be associated with poorer cognitive function later in life overall, and particularly among defenders and goalkeepers. It is notable that the latter group rarely, if ever, head the ball, but do suffer from concussions. Taken overall, these results are consistent with an increased risk of poorer cognitive function resulting from more severe impacts. A previous systematic review of the literature suggested that Peak Linear Acceleration (PLA) varied across studies, but was higher overall for other impacts to the head - such as head-to -head collisions - than exposure to heading, in males^26^. Consistently, impacts leading to neurological symptoms, thus classifiable as concussions, were found at the top 1% by magnitude of PLA among in a study among military and civil volunteers^35^.

Overall, these findings suggest that there is little or no association between heading the ball and reduced cognitive function. This is consistent with the findings of some previous studies^10,13,14^, recently summarised in a systematic review^15^. Previous cross-sectional studies that found an association between heading the ball and cognitive function^7,11,12^ used various questionnaires to collect data on player exposure characteristics including playing position, with some of the studies also collecting data on the number of headers^11,12^. It is unclear, however, how well these were able to capture gaps in playing history due to injury or unemployment. None of these studies attempted to validate their data against the playing career. The only study which relied on a sports database for number of matches during their career was by Rodrigues et al^13^. Here, a validation of the self-reported number of headers per game against observations of currently active players during 42 games was carried out, and this was reassuring, although the results may not apply to recall of subjects who played many decades in the past. The study found no association between exposure to heading and performance on neuropsychological tests^13^.

In our study, we used a questionnaire that was developed, piloted and validated in a group of former professional players. Further validation by comparing the participants recall against available historical records showed that only 6% of the participants incorrectly reported the duration of their playing career and the order of the clubs they had played for (manuscript in preparation). There is a need to also validate the reliability of recall of historic data on the number of headers per game or training session, and it would be advantageous to standardise the relevant questionnaire tools. Moreover, in our study we used quantitative exposure estimates on the cumulative number of head impacts experienced during training and play, derived from empirical exposure modelling approaches. This allowed the assignment of exposure on a group-based level, similar to when implementing Job Exposure Matrices^36^. Use of group-based exposure assessment approaches limit the potential for bias on the observed associations between exposure and health outcomes due to the introduction of “Berkson error”^37^. Our regression model estimates, pending some further validation, could be used in other studies, including those already conducted in Scotland^5^, Sweden^8^ and France^9^, if data on playing position, level of play, and duration were available.

Our findings also suggest that exposure to head traumas more severe than heading a ball could be associated with poorer cognitive function later in life. Head-to-head collisions and other blows to the head, were found to be associated with poorer cognitive function, although this was among forwards only, and the possibility of a chance finding (because of multiple subgroup analysis) cannot be ruled out. Soccer-related concussions, on the other hand, were associated with an increased risk of poor cognitive function in the overall sample. Each additional soccer-related concussion was associated with a decrease in cognitive function of about 0.01 SD below the mean PACC score compared to former players of the same age. This is a novel finding, despite the relevant information being collected in some of the previous studies^11^, but not all^12^. This association is consistent with results from other sports such as boxing or American football, where concussion was related to poorer cognitive function^1,38^. Importantly, this finding suggests that the previous studies based on external comparison which found a higher incidence or mortality from dementia among soccer players compared to the general population^5,7–9^, might – at least partially – be explained by exposures to soccer-related concussions, rather than heading the ball. If further confirmed, this might have important implications in informing the regulation of the game to prevent increasing the risk of dementia. On the other hand, it is important to note that the size of the observed effect per concussion was very small (0.01 SD), and its clinical significance is therefore unclear. Interestingly, the age-adjusted effect estimate was comparable to that previously found among rugby players (• = -0.01, 95% C.I. -0.04, 0.03).

The main limitation of this study is the cross-sectional design, and therefore the potential for recall bias. This was minimized by using appropriate modelled estimation of the exposure to heading, other head impacts, and the BRAIN-Q tool to assess history of concussion^28^. Selection bias may have arisen from a suboptimal response rate, and because participants with more concussions appear to have been more likely to participate. However, this will not necessarily bias the main study analyses as they involve internal comparisons. Conversely, assuming that our sample may be also biased toward participants with better cognitive function, this may lead to an overestimation of any association between concussion and cognitive function, but any such bias is likely to be small, and unlikely to account entirely for the association found. Unfortunately, it was not possible to estimate pre-morbid intelligence in this study. It is also not possible to exclude that the association between concussion and cognitive function may be modified by some genetic trait, for example, apolipoprotein E (*APOE*) genotype. Finally, no data on depression were collected.

In conclusion, this study does not support an association between heading a ball and poorer cognitive function later in life among former male professional soccer players. However, it does suggest an association between concussion and poorer cognitive function in the same group. This result is consistent with previous studies assessing external comparisons on incidence/mortality of dementia among soccer players and population controls. If further replicated and confirmed, this finding will help to shape policies for reducing the risk of occupationally-related dementia among soccer players.

## Data Availability

Data sharing Pseudonymised data may be made available on request to external researchers for additional analyses meeting the scientific and ethical standards under which the data was collected, and consistent with the overall aims of the project.
If such requests are approved, these additional analyses would be done in collaboration with the study team, and only after approval of the Independent Oversight Committee for the study, and after additional ethical approved, if required.

## Data sharing

Pseudonymised data may be made available on request to external researchers for additional analyses meeting the scientific and ethical standards under which the data was collected, and consistent with the overall aims of the project.

If such requests are approved, these additional analyses would be done in collaboration with the study team, and only after approval of the Independent Oversight Committee for the study, and after additional ethical approved, if required.

### Acknowledgements

We are grateful to all the HEADING study participants for the time, interest, and commitment they have shown in contributing to the data collection. Thank you to the Football Association (FA) for their continued support with recruitment. We would like to thank Prof Carol Brayne who has provided very valuable input on each of the phases of the HEADING study by chairing the BRAIN and HEADING study Independent Oversight Committee (IOC). Thanks also to the IOC Members for their invaluable advice and guidance throughout: Bill Treadwell, Simon Jones, Dr. Collette Griffin, Professor Sinead Langan, Tim Lindsay, Hannah Wilson and Tim Stevens

We also would like to thank Dr Hannah Whiteman, Dr Alexandra Anderson, Dr Heiner Grosskurth, Dr Jennifer Nicholas, Mr James Barr, Dr Peter Wright and the other collaborators at London School of Hygiene and Tropical Medicine.

## Availability of data and material

Ms Seghezzo, Dr Williamson and Prof Pearce had full access to all of the data in the study and take responsibility for the integrity of the data and the accuracy of the data analysis. Dr Gallo declares that this manuscript is honest, accurate, and transparent account of the study being reported; that no important aspects of the study have been omitted. All co-authors had full access to the data, and can take responsibility for the integrity of the data and the accuracy of the data analysis.

## Authors contributions

Study concept and design: V Gallo, N Pearce, D McElvenny

Analysis and interpretation of data: G Seghezzo, E Williamsons, V Gallo, N Pearce, D McElvenny, I Basinas, S Kemp

Drafting of the manuscript:V Gallo

Data collection: S Mian, D Davoren, D Pearce, Y van der Hoecke

Critical revision of the manuscript for important intellectual content: G Seghezzo, I Basinas, E Williamson, Y van der Hoecke, S Kemp, S Langan, H Zetterberg, JW Cherrie, DM McElvenny, N Pearce

Conflict of interest statement: HZ has served at scientific advisory boards and/or as a consultant for Abbvie, Acumen, Alector, Alzinova, ALZPath, Amylyx, Annexon, Apellis, Artery Therapeutics, AZTherapies, Cognito Therapeutics, CogRx, Denali, Eisai, Merry Life, Nervgen, Novo Nordisk, Optoceutics, Passage Bio, Pinteon Therapeutics, Prothena, Red Abbey Labs, reMYND, Roche, Samumed, Siemens Healthineers, Triplet Therapeutics, and Wave, has given lectures in symposia sponsored by Alzecure, Biogen, Cellectricon, Fujirebio, Lilly, Novo Nordisk, and Roche, and is a co-founder of Brain Biomarker Solutions in Gothenburg AB (BBS), which is a part of the GU Ventures Incubator Program (outside submitted work). HZ is a Wallenberg Scholar and a Distinguished Professor at the Swedish Research Council supported by grants from the Swedish Research Council (#2023-00356; #2022-01018 and #2019-02397), the European Union’s Horizon Europe research and innovation programme under grant agreement No 101053962, Swedish State Support for Clinical Research (#ALFGBG-71320), the Alzheimer Drug Discovery Foundation (ADDF), USA (#201809-2016862), the AD Strategic Fund and the Alzheimer’s Association (#ADSF-21-831376-C, #ADSF-21-831381-C, #ADSF-21-831377-C, and #ADSF-24-1284328-C), the Bluefield Project, Cure Alzheimer’s Fund, the Olav Thon Foundation, the Erling-Persson Family Foundation, Stiftelsen för Gamla Tjänarinnor, Hjärnfonden, Sweden (#FO2022-0270), the European Union’s Horizon 2020 research and innovation programme under the Marie Skłodowska-Curie grant agreement No 860197 (MIRIADE), the European Union Joint Programme – Neurodegenerative Disease Research (JPND2021-00694), the National Institute for Health and Care Research University College London Hospitals Biomedical Research Centre, and the UK Dementia Research Institute at UCL (UKDRI-1003).

---

**Supplementary Table 1:** Characteristics of study participants by categories of soccer-related concussions

|  | No concussion | Low concussion<br>(1-2) | High concussion<br>(3+) | All |
| --- | --- | --- | --- | --- |
|  | N= 33 | N= 52 | N= 55 | N= 142 |
| **Number of Soccer-Related Concussions, median (p25, p75)<sup>1</sup>** | 0 | 1 (1, 2) | 5 (3, 10) | 2 (1, 4) |
| **Age (years), median (p25, p75)** | 66 (60, 75) | 65 (60, 71.5) | 59 (55, 67) | 63 (58, 71) |
| **Educations (years), median (p25, p75)<sup>1</sup>** | 11 (10, 12) | 11 (10, 12) | 11 (10, 14) | 11 (10, 12.5) |
| **Education, N (%)** |  |  |  |  |
| *Primary School* | 7 (38.1) | 6 (28.6) | 7 (33.3) | 21 (14.8) |
| *GCSE or equivalent* | 16 (20.3) | 37 (46.8) | 26 (32.9) | 80 (56.3) |
| *A-Level or above* | 9 (22.5) | 9 (22.5) | 22 (55) | 41 (28.9) |
| **Ethnicity, N (%)** |  |  |  |  |
| *White* | 29 (23.2%) | 45 (36%) | 51 (40.9%) | 126 (88.7%) |
| *Black* | 4 (30.8%) | 6 (46.2%) | 3 (23.1%) | 14 (9.9%) |
| *Mixed* | 0 | 1 (50%) | 1 (50%) | 2 (1.4%) |
| **Smoking, N (%) <sup>2</sup>** |  |  |  |  |
| *Never smoker* | 24 (20.5) | 39 (36.8) | 42 (39.6) | 119 (83.8) |
| *Former smoker* | 5 (33.3) | 3 (20) | 7 (46.7) | 15 (10.6) |
| *Current smoker* | 4 (50) | 3 (37.5) | 1 (12.5) | 8 (5.6) |
| **Alcohol, N (%) <sup>2</sup>** |  |  |  |  |
| *Never drinker* | 6 (26.1) | 10 (43.5) | 7 (30.4) | 24 (16.9) |
| *Former drinker* | 1 (25) | 1 (25) | 2 (50) | 4 (2.8) |
| *Current drinker* | 26 (23) | 41 (36.3) | 46 (40.7) | 114 (80.3) |
| **Hypertension, N (%)** |  |  |  |  |
| *Yes* | 7 (19.4) | 16 (44.4) | 13 (36.1) | 37 (26.1) |
| *No* | 26 (25) | 36 (34.6) | 42 (40.4) | 105 (73.9) |
| **Diabetes, N (%)** |  |  |  |  |
| *Yes* | 0 | 3 (75) | 1 (25) | 5 (3.5) |
| *No* | 33 (24.3) | 49 (36) | 54 (39.7) | 137 (96.5)) |
| **Position played, N (%) <sup>3</sup>** |  |  |  |  |
| *Defence* | 8 (17.8) | 16 (35.6) | 21 (46.7) | 46 (35.1)) |
| *Forward* | 7 (29.2) | 8 (33.3) | 9 (37.5) | 24 (18.3) |
| *Goalkeeper* | 0 | 3 (75) | 1 (25) | 4 (3.1) |
| *Midfield* | 6 (22.2) | 11 (40.7) | 10 (37.0) | 27 (20.6) |
| *Unknown* | 8 (22.5) | 49 (38) | 51 (40) | 30 (22.9) |
| **Length of career (years), mean (SD) <sup>3</sup>** | 15.9 (6.03) | 19.2 (5.8) | 16.7 (5.8) | 17.5 (5.9) |
| **Study centre, N (%) <sup>1</sup>** |  |  |  |  |
| *ISEH, London* | 2 (22.2) | 3 (33.3) | 4 (44.4) | 9 (6.3) |
| *Manchester* | 3 (21.4) | 5 (35.7) | 6 (42.9) | 14 (9.9) |
| *Home* | 3 (15) | 10 (50) | 7 (35) | 20 (14.1) |
| *Remote* | 25 (25.3) | 34 (34.3) | 38 (38.4) | 99 (69.7) |
| **MMSE score below 25, N (%)** | 5 (21.7%) | 7 (30.4%) | 10 (43.5%) | 23 (16.2) |

1. Two missing values; 2. smoking and alcohol categorised into ever/never in the multivariable model; 3. eleven missing values

**Supplementary Table 2:** Median (p25-p75) scores for the individual cognitive function tests, by concussion/quartiles of cumulative

|  | N | PACC score,<br>median (p25,<br>p75) | MMSE,<br>median<br>(p25, p75) | FNAME<br>score,<br>median<br>(p25, p75) | LMDR,<br>median<br>(p25, p75) | Digit<br>Symbol<br>Substitutio<br>n, median<br>(p25, p75) |
| --- | --- | --- | --- | --- | --- | --- |
| **All Ages** |  |  |  |  |  |  |
| No Concussions | 32 | -0.13 (-0.57, 0.54) | 29 (27, 30) | 40 (17, 64) | 7 (4, 11) | 43 (34, 51) |
| Low Concussions (1-2) | 49 | 0.09 (-0.18, 0.43) | 29 (28, 30) | 52 (31, 66) | 8 (6, 11) | 48 (41, 54) |
| High Concussions (3+) | 52 | 0.20 (-0.18, 0.62) | 29 (27, 30) | 56 (39, 67) | 9 (6, 11) | 47 (36, 55) |
| Heading<sup>1</sup> 1<sup>st</sup> quartile | 28 | 0.22 (-0.30, 0.63) | 29 (29, 30) | 50 (38, 68) | 8 (5, 12) | 49 (40, 55) |
| Heading<sup>1</sup> 2<sup>nd</sup> quartile | 30 | 0.02 (-0.43, 0.48) | 29 (28, 30) | 48 (31, 66) | 8 (5, 11) | 45 (34, 55) |
| Heading<sup>1</sup> 3<sup>rd</sup> quartile | 32 | 0.24 (-0.14, 0.55) | 29 (28, 30) | 56 (35, 66) | 9 (6, 11) | 48 (41, 55) |
| Heading<sup>1</sup> 4<sup>th</sup> quartile | 23 | -0.11 (-0.49, 0.49) | 27 (25, 29) | 45 (28, 65) | 8 (5, 10) | 45 (36, 53) |

**Supplementary Table 3:** age-adjusted and fully adjusted beta coefficinets coming from linear regression models investigating the association between self-reported heading, impacts to the head, and soccer-related concussion with the PACC score among former professional soccer players

|  | N | Age-adjusted Coefficient<br>(95% CI) | Fully adjusted<br>Coefficient (95% CI) | Mutually adjusted<br>Coefficients^ (95% C.I.) |
| --- | --- | --- | --- | --- |
| **Heading <sup>1</sup>** |  |  |  |  |
| 1<sup>st</sup> quartile | 50 | Reference | Reference | Reference |
| 2<sup>nd</sup> quartile | 52 | -0.04 (-0.29, 0.21) | -0.01 (-0.23, 0.24) | -0.03 (-0.21, 0.28) |
| 3<sup>rd</sup> quartile | 49 | -0.02 (-0.28, 0.23) | -0.02 (-0.27, 0.22) | -0.02 (-0.27, 0.23) |
| 4<sup>th</sup> quartile | 48 | -0.24 (-0.55, 0.06) | -0.20 (-0.51, 0.11) | -0.18 (-0.49, 0.13) |
| p-trend |  | 0.304 | 0.376 | 0.945 |
| Number of headers (continuous per 1,000 episodes) | 199 | -0.003 (-0.005, -0.0006) | -0.003 (-0.005, -0.0004) | -0.003 (-0.005, -0.0005) |
| **Other impacts to the head <sup>1</sup>** |  |  |  |  |
| 1<sup>st</sup> quartile | 34 | Reference | Reference | Reference |
| 2<sup>nd</sup> quartile | 59 | 0.03 (-0.32, 0.38) | -0.002 (-0.31, 0.31) | 0.02 (-0.32, 0.32) |
| 3<sup>rd</sup> quartile | 52 | -0.08 (-0.51, 0.34) | -0.10 (-0.47, 0.27) | -0.08 (-0.48, 0.32) |
| 4<sup>th</sup> quartile | 54 | 0.01 (-0.31, 0.32) | -0.07 (-0.37, 0.22) | 0.03 (-0.34, 0.27) |
| p-trend |  | 0.887 | 0.873 | 0.945 |
| Number of other impacts to the head (continuous per 1,000 episodes) | 199 | 0.003 (-0.02, 0.03) | -0.0003 (-0.02, 0.02) | 0.007 (-0.01, 0.03) |
| **Soccer-related Concussions** |  |  |  |  |
| No Concussions | 28 | Reference | Reference | Reference |
| Low Concussions (1-2) | 41 | 0.07 (-0.18, 0.32) | 0.09 (-0.16, 0.34) | 0.09 (-0.17, 0.34) |
| High Concussions (3+) | 47 | -0.08 (-0.33, 0.17) | -0.09 (-0.32, 0.01) | -0.06 (-0.30, 0.19) |
| p-trend |  | 0.410 | 0.274 | 0.945 |
| Number of concussions (continuous) | 199 | -0.01 (-0.03, -0.001) | -0.01 (-0.02, -0.002) | -0.01 (-0.02, 0.00006) |
| **Length of career, years** | 199 | -0.004 (-0.02, 0.01) | -0.003 (-0.02, 0.01) | -0.003 (-0.02, 0.010) |

1. number of headers and other impacts to the head estimated from playing history <sup>19</sup>; 2. Mutually adjusted models include the three categorical exposure metrics (quartiles of heading and of other impacts to the head) and categories of concussion or the three continuous exposure metrics (number of headers, number of other impacts to the head, and number of concussions) in the same model. Length of career is adjusted for the three continuous metrics.

**Supplementary Table 4:** fully adjusted beta coefficients coming from linear regression models investigating the association between estimated training-related and in game-related heading episodes with the PACC score among former professional soccer players

|  | N | Fully-adjusted<br>Coefficient (95% CI)<br>Training | N | Fully-adjusted<br>Coefficient (95% CI)<br>During game |
| --- | --- | --- | --- | --- |
| **Heading <sup>1</sup>** |  |  |  |  |
| 1<sup>st</sup> quartile | 50 | Reference | 52 | Reference |
| 2<sup>nd</sup> quartile | 50 | -0.14 (-0.47, 0.19) | 50 | -0.13 (-0.40, 0.15) |
| 3<sup>rd</sup> quartile | 51 | -0.14 (-0.42, 0.14) | 50 | -0.03 (-0.29, 0.23) |
| 4<sup>th</sup> quartile | 48 | -0.16 (-0.48, 0.16) | 47 | -0.07 (-0.36, 0.22) |
|  | p-trend | 0.720 | p-trend | 0.830 |

1. number of headers and other impacts to the head estimated from playing history <sup>19</sup>, the number of players in each quartiles are average across imputations

**Supplementary Table 5:** Sensitivity analysis of the age-adjusted and fully adjusted beta coefficinets coming from linear regression models investigating the association between estimated heders, impacts to the head, and soccer-related concussion with the PACC score among former porfessional soccer players on non-imputed data

|  | N | Age-adjusted Coefficient<br>(95% CI) | N | Fully adjusted<br>Coefficient (95% CI) | N | Mutually adjusted<br>Coefficients^ (95% C.I.) |
| --- | --- | --- | --- | --- | --- | --- |
| **Heading \*** |  |  |  |  |  |  |
| 1<sup>st</sup> quartile | 24 | Reference | 24 | Reference | 23 | Reference |
| 2<sup>nd</sup> quartile | 29 | -0.002 (-0.31, 0.31) | 29 | -0.03 (-0.33, 0.27) | 29 | -0.11 (-0.43, 0.22) |
| 3<sup>rd</sup> quartile | 29 | 0.10 (-0.21, 0.40) | 29 | 0.01 (-0.29, 0.31) | 29 | -0.06 (-0.38, 0.27) |
| 4<sup>th</sup> quartile | 18 | -0.05 (-0.39, 0.30) | 18 | -0.03 (-0.39, 0.34) | 18 | -0.008 (-0.39, 0.38) |
| p-trend |  | 0.841 | p-trend | 0.987 | p-trend | 0.882 |
| Number of headers (continuous, per 1,000 episodes) | 102 | -0.008 (-0.01, -0.001) | 102 | -0.004 (-0.01, -0.004) | 100 | -0.003 (-0.01, 0.005) |
| **Other impacts to the head\*** |  |  |  |  |  |  |
| 1<sup>st</sup> quartile | 29 | Reference | 29 | Reference | 14 | Reference |
| 2<sup>nd</sup> quartile | 28 | 0.01 (-0.31, 0.33) | 28 | 0.07 (-0.22, 0.37) | 27 | 0.16 (-0.22, 0.55) |
| 3<sup>rd</sup> quartile | 30 | 0.06 (-0.25, 0.38) | 30 | 0.05 (-0.25, 0.34) | 29 | 0.14 (-0.21, 0.50) |
| 4<sup>th</sup> quartile | 30 | -0.19 (-0.51, 0.13) | 30 | -0.27 (-0.57, 0.3) | 29 | -0.17 (-0.57, 0.22) |
| p-trend |  | 0.407 | p-trend | 0.09 |  | 0.07 |
| Number of other impacts to the head (continuous) | 103 | -0.0015 (-0.004, 0.001) | 103 | -0.0017 (-0.004, 0.0009) | 100 | -0.0013 (-0.004, 0.001) |
| **Soccer-related Concussions** |  |  |  |  |  |  |
| No Concussions | 28 | Reference | 26 | Reference | 24 | Reference |
| Low Concussions (1-2) | 41 | 0.19 (-0.10, 0.49) | 40 | 0.15 (-0.13, 0.42) | 26 | 0.13 (-0.15, 0.41) |
| High Concussions (3+) | 47 | -0.05 (-0.24, 0.35) | 42 | -0.04 (-0.28, 0.27) | 39 | 0.08 (-0.20, 0.36) |
| p-trend |  | 0.363 | p-trend | 0.404 | p-trend | 0.65 |
| Number of concussions (continuous) | 116 | -0.01 (-0.03, -0.004) | 109 | -0.02 (-0.03, -0.004) | 100 | -0.02 (-0.04, 0.006) |
| **Length of career, years** | 110 | -0.002 (-0.02, 0.02) | 110 | -0.002 (-0.02, 0.02) | 101 | -0.003 (-0.02, 0.02) |

